# Knowledge and Risk Perception of The Novel Coronavirus Disease 2019 Among Adult Nigerians: A Cross-Sectional Study

**DOI:** 10.1101/2020.08.23.20180141

**Authors:** Erick Wesley Hedima, Samuel Adeyemi Michael, Emmanuel Agada David

## Abstract

COVID-19 caused by severe acute respiratory syndrome coronavirus 2 (SARS*-*CoV*-*2) is a highly infectious disease declared a pandemic by the World Health Organization. The Knowledge and risk perception in the adult population may influence adherence to safety guidelines.

**Objective:** To assess the knowledge, preventive measures and risk perception of adult Nigerians regarding COVID-19.

**Methods:** We conducted an online cross-sectional survey in which five hundred and ten (510) adult participants consented and filled the questionnaire. The questionnaire is divided in to four sections: 1) socio-demographic characteristics of the participants, 2) assessment of knowledge, 3) risk perception and the 4) preventive measures.

**Results:** Of the 510 respondents, 95.9% claimed knowledge of COVID-19, through the traditional media (55.3%), and social media (41%), while only 3.7% got informed through health officials. Level of education (*P*=0.0001), income status (*P*<0.00001) and being a healthcare worker (*P*=0.002) were significantly associated with a good knowledge of COVID-19. Overall Risk perception was high (median score of 4 out of 5). Risk perception was significantly high among the female participants (*P*=0.04), young adult (*P*=0.039) and healthcare providers (*P*=0.001), while knowledge of preventive measures like avoiding to eat outside the home (*P*=0.001), traveling to high risk areas (*P*=0.017), wearing face mask (*P*=0.01) and eating balanced diet (*P*=0.014) were significant across gender.

**Conclusion:** Most participants demonstrated good knowledge of COVID-19 and its preventive measures, while risk perception was higher among healthcare workers. Findings from this survey could guide information campaigns by public health authorities, clinicians, and the media.

## Introduction

The latest threat to global health is the ongoing outbreak of the respiratory disease that was recently given the name Coronavirus Disease 2019 (Covid-19). Covid-19 was recognized in December 2019^1^. The highly contagious severe acute respiratory syndrome coronavirus (SARS-CoV-2) which emanated from China and has since become a global public health emergency^2^. In severe cases, the virus causes fatal pneumonia similar to that caused by SARS and Middle East respiratory syndrome coronavirus (MERS-CoV), which had emerged in the past years sporadically in countries^3^. The course of the Covid-19 epidemic will likely be strongly impacted by how the population behaves, which in turn is influenced by what people know and believe about this disease^4^. A particular concern in this regard is the spread of misinformation about COVID-19 on social media. This has led the WHO to host a page with “myth busters” on the world body’s website and engage in discussions on the social media^5^. There is a great concern by the World Health Organization that COVID-19 could take time to eliminated, and that the rate at which the infection is spreading across the world calls for rapid assessment of the population’s knowledge and perceptions of this infection^6,7^.

This work is aimed to assess the knowledge, the preventive measures and risk perception of adult Nigerian population regarding the novel coronavirus disease 2019 (COVID-19).

## Methods

### Study design and settings

This was a web-based cross-sectional survey among adult Nigerian population.

### Study tools

The survey questionnaire was adopted from other studies^8,9^. It covered the socio-demographic characteristics, knowledge regarding COVID-19 and its preventive measures and perceived risk about the disease.

### Pilot study

A pilot study was conducted to assess the reliability of the questionnaire before its use. The questionnaire was pretested on 20 participants who were excluded later from the main study. Participants completed the perceived risk scale (Cronbach’s α = 0.82) which had 8 survey-items (5-point Likert scale, from strongly disagree to strongly agree).

### Data collection

An online survey portal, Google Form was created, and adult participants were invited to complete and submit the form via WhatsApp, Facebook and Twitter social media sites. The process of calling participants to share in the survey was conducted through snowball sampling techniques^10^. Participants continued to spread and were expected to cover the entire six regions of the country. The study was conducted from May to July, 2020 among Nigerian adults.

### Sampling

The sample size was determined using the Epi Info 7.0 software (Centers for Disease Control and Prevention, Atlanta, USA)^11^. As there were few similar studies related to coronavirus disease in Nigeria, the calculations were based on the assumption that the probability of having good knowledge on preventive measures against coronavirus disease was 50.0%. Using the margin of error of 5%, a design effect of 1.0, and the confidence interval set at 95%^12^. The calculated sample size was 384 participants. The survey portal was closed, and interviews stopped at the end of the day when the number of participants exceeded the sample size, i.e. at the end of the fifth week. The online questionnaire was designed in such a way as to allow for only one response per participant.

### Statistical Analysis

Participant’s responses were analyzed using the Statistical Package for Social Sciences (SPSS) version 25.0^13^. Descriptive statistics were used to summarize data on socio-demographic characteristics, infection prevention and control measures against the novel coronavirus by participants and responses to questions concerning knowledge and risk perceptions towards the new coronavirus. Continuous variables were presented as mean and median, depending on items’ distribution, while categorical variables were reported as frequencies (n) and proportions (%). Each item on the knowledge of COVID-19 was assigned ‘1 mark’ for one correct response and a ‘higher mark’ for a wrong response, thereby making participants with good knowledge to have a lower score. The total score for individual respondent was computed and categorized as ‘good (≤ 25)’, ‘fair (26 −35)’, ‘poor (36 - 45)’ and ‘very poor (≥ 46)’ knowledge, depending on the cumulative scores. Chi-square test was performed to determine association between socio-demographic characteristics and knowledge as well as infection prevention and control measures. A post hoc test was carried out after a significant Chi square test to identify where the difference in knowledge of the disease really lies. *Kruskal Wallis* test with post-hoc was used to assess difference in the risk perception across sociodemographic characteristics. P-value < 0.05 was considered statistically significant at 95% confidence interval (CI).

### Ethical Considerations

This study was approved by the Ethics Committee of Gombe State University. Participants’ anonymity and confidentiality were ensured. A Participant information sheet was served and an informed consent was obtained before the participant answered the questionnaire.

## Results

Five hundred and ten (510) persons from the 6 regions of Nigeria completed the survey. **Table 1** shows the socio-demographic characteristics of the studied participants. More than two thirds (66.9%) were males. More than half the participants (53.5%) aged 26 to <35 years, less than a quarter aged 18 to less than 25 (16.3%), aged 36 to less than 45 (14%) and aged 46 to ≤55, whereas only 3.7% aged 55 and above. Most of the participant reside in the north east (37.6%). More than half were university graduates (56.5%), 23.7% had Master’s Degree, 3.7% had Doctorate degrees respectively. The monthly income of a large proportion of the participants 38.6% was more than 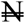 110,000. More than half of the participants (59.4%) were not healthcare workers. Of the 40.6% healthcare workers who responded, 19.2% were pharmacists, 2.2% were physicians, while only 2.5% were nurses.

**Table 1:** Socio-demographic characteristics of the participants (n = 510)

| **Socio-demographic characteristics** | **n (%)** |
| --- | --- |
| **Gender** |  |
| Male | 341 (66.9) |
| Female | 169 (33.1) |
| **Age** |  |
| 18 – 25 | 83 (16.3) |
| 26 – 35 | 273 (53.5) |
| 36 – 45 | 92 (18.0) |
| 46 – 55 | 43 (8.4) |
| 55+ | 19 (3.7) |
| **Current level of education** |  |
| Less than senior school certificate | 2 (0.4) |
| Senior school certificate | 14 (2.7) |
| Diploma | 35 (6.9) |
| Bachelor's degree | 288 (56.5) |
| Master's degree | 121 (23.7) |
| Professional degree | 31 (6.1) |
| Doctorate | 19 (3.7) |
| **Region of Residence** |  |
| South East | 28 (5.5) |
| North Central | 128 (25.1) |
| South South | 15 (2.9) |
| North West | 52 (10.2) |
| South West | 95 (18.6) |
| North East | 192 (37.6) |
| **Monthly income (Naira)** |  |
| <30, 000 | 151 (29.6) |
| 30, 000 – 59, 000 | 73 (14.3) |
| 60, 000 – 89, 000 | 47 (9.2) |
| 90, 000 – 109, 000 | 42 (8.2) |
| ≥110, 000 | 197 (38.6) |
| **Are you a healthcare worker?** |  |
| No | 303 (59.4) |
| Nurse | 13 (2.5) |
| Physician | 11 (2.2) |
| Community health worker | 11 (2.2) |
| Pharmacist | 98 (19.2) |
| Med. Laboratory Scientist | 22 (4.3) |
| Other healthcare worker | 52 (10.2) |

Almost all of the participants (95.9%) claimed they were aware of the novel coronavirus. Majority (55.3%) of the participants were aware of COVID-19 mostly through the media (TV/Radio/Bill boards/Newspapers). Only a small percentage (3.7%) were aware through health officials. Nearly half of the participants (48%) had a good knowledge of the disease; 34.9% had a fair knowledge, 13.9% had a poor knowledge and 3.1% had a very poor knowledge of the disease.

**Figure 1.**
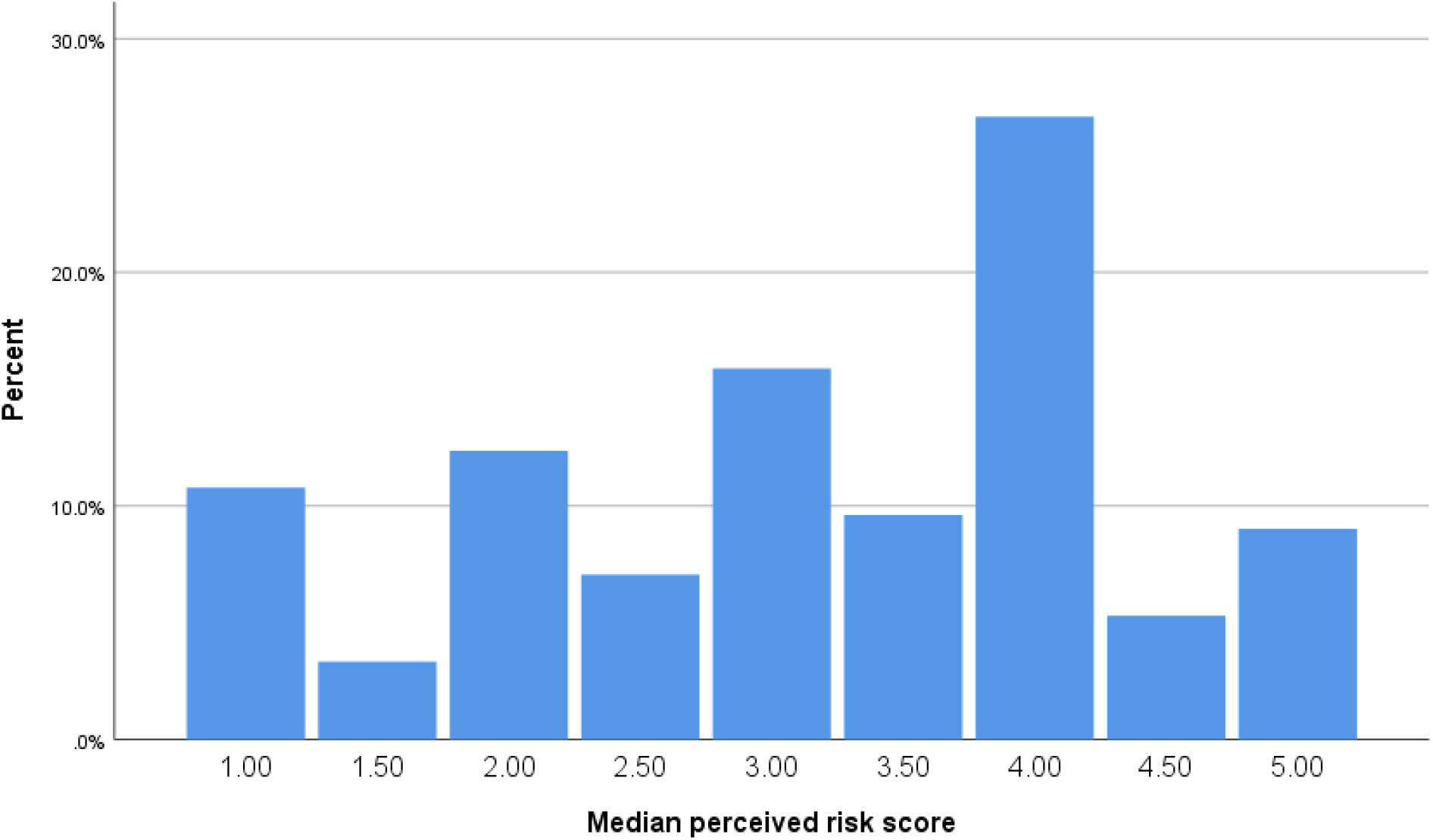
Distribution of risk perception. The median risk perception score was 4.0 out of a total of 5 (Range = 4; Fig 1).

The relationship between socio-demographic characteristics and knowledge about COVID-19 is demonstrated in **Table 2**. Good knowledge score of COVID-19 was significantly related to higher level of education (*p*<0.01) and monthly income (*p*<0.001). Being a healthcare worker also influence the knowledge about COVID-19 (*p*=0.002). When asked *“If you have a fever or cough and recently came in contact with someone who is confirmed to be positive for COVID-19, what action will you take?* 61% of the participants responded with the recommended care-seeking option of staying home and contacting their healthcare system. More than a quarter (33.3%) of the participants stated they would delay care-seeking by self-isolation while a small percent (0.6%) of the participants would rather attend the hospital emergency department and 2% of the participants would rather rest more than usual and if symptom persists, they take a public transport to their primary care provider.

**Table 2:** Relationship between socio-demographic characteristics of the participants and their knowledge scores about COVID-19 (n = 510)

| Socio-demographic characteristics | Knowledge category |  |  |  |  | P-value |
| --- | --- | --- | --- | --- | --- | --- |
|  | Good | Fair | Poor | Very poor | Total |  |
| **Gender** |  |  |  |  |  |  |
| Male | 170 (69.4) | 117 (65.7) | 43 (60.6) | 11(68.8) | 341 (66.9) | 0.55 |
| Female | 75 (30.6) | 61 (34.3) | 28 (39.4) | 5 (31.3) | 169 (33.1) |  |
| **Age** |  |  |  |  |  |  |
| 18 – 25 | 35 (14.3) | 33 (18.5) | 15 (21.1) | 0 (0) | 83 (16.3) | 0.81 |
| 26 – 35 | 132 (53.9) | 96 (53.9) | 36 (50.7) | 9 (56.3) | 273 (53.5) |  |
| 36 – 45 | 47 (19.2) | 30 (16.9) | 11 (15.5) | 4 (25) | 92 (18) |  |
| 46 – 55 | 20 (8.2) | 14 (7.9) | 7 (9.9) | 2 (12.5) | 43 (8.4) |  |
| 55+ | 11 (4.5) | 5 (2.8) | 2 (2.8) | 1 (6.3) | 19 (3.7) |  |
| **Current level of education** |  |  |  |  |  |  |
| Less than SSCE | 0 (0) | 0 (0.0) | 1 (1.4) | 1 (6.3) | 2 (0.4) | 0.003* |
| SSCE | 6 (2.4) | 6 (3.4) | 0 (0.0) | 2 (12.5) | 14 (2.7) |  |
| Diploma | 11 (4.5) | 11 (6.2) | 9 (12.7) | 4 (25.0) | 35 (6.9) |  |
| Bachelor's degree | 134 (54.7) | 109 (61.2) | 40 (56.3) | 5 (31.3) | 288 (56.5) |  |
| Master's degree | 68 (27.8) | 34 (19.1) | 16 (22.5) | 3 (18.8) | 121(23.7) |  |
| Professional degree | 14 (5.7) | 11(6.2) | 5 (7.0) | 1 (6.3) | 31 (6.1) |  |
| Doctorate | 12 (4.9) | 7 (3.9) | 0 (0.0) | 0 (0) | 19 (3.7) |  |
| **Region of residence** |  |  |  |  |  |  |
| South East | 10 (4.1) | 14 (7.9) | 3 (4.2) | 1 (5.5) | 28 (5.5) | 0.42 |
| North Central | 57 (23.3) | 47 (26.4) | 18 (25.4) | 6 (25.1) | 128 (25.1) |  |
| South South | 10 (4.1) | 4 (2.2) | 1 (1.4) | 0 (2.9) | 15 (2.9) |  |
| North West | 24 (9.8) | 15 (8.4) | 13 (18.3) | 0 (10.2) | 52 (10.2) |  |
| South West | 50 (20.4) | 33 (18.5) | 10 (14.1) | 2 (18.6) | 95 (18.6) |  |
| North East | 94 (38.4) | 65 (36.5) | 26 (36.6) | 7 (37.6) | 192 (37.6) |  |
| **Average Monthly income (Naira)** |  |  |  |  |  |  |
| <30, 000 | 57 (23.3) | 55 (30.9) | 30 (42.3) | 9 (56.3) | 151 (29.6) | <0.001* |
| 30, 000 – 59, 000 | 30 (12.2) | 29 (16.3) | 12 (16.9) | 2 (12.5) | 73 (14.3) |  |
| 60, 000 – 89, 000 | 22 (9.0) | 20 (11.2) | 3.0 (4.2) | 2 (12.5) | 47 (9.2) |  |
|  |  |  |  |  | 42 (8.2) |  |
| 90, 000 – 109, 000 | 15 (6.1) | 15 (8.4) | 11 (15.5) | 1 (6.3) |  |  |
| ≥110, 000 | 121 (49.4) | 59 (33.1) | 15 (21.1) | 2 (12.5) | 197 (38.6) |  |
| **Are you a HCW?** |  |  |  |  |  |  |
| No | 133 (54.3) | 115 (64.6) | 49 (69) | 6 (59.4) | 303 (59.4) | 0.002* |
| Nurse | 6 (2.4) | 5 (2.8) | 1 (1.4) | 1 (37.5) | 13 (2.5) |  |
| Physician | 7 (2.9) | 2 (1.1) | 2 (2.8) | 0 (00.0) | 11 (2.2) |  |
| CHW | 3 (1.2) | 6 (3.4) | 1 (1.4) | 1 (6.3) | 11 (2.2) |  |
| Pharmacist | 57 (23.3) | 32 (18.0) | 9 (12.7) | 0 (0.0) | 98 (19.2) |  |
| MLS | 16 (6.5) | 2 (1.1) | 3 (4.2) | 1 (6.3) | 22 (4.3) |  |
| Other healthcare worker | 23 (9.4) | 16 (9.0) | 6 (8.5) | 7 (43.8) | 52 (10.2) |  |

**KEY:** SSCE= Senior School Certificate Examination; HCW= Healthcare worker; CHW= Community health worker; MLS= Medical Lab Scientist; \*Statistically significant at $p < 0.05$ , 95% CI

However, a post-hoc test was carried out to identify the association between knowledge of the novel coronavirus and sociodemographic characteristics in those that were significant using chi-square test in which lower level of formal education (*p*=0.0001) and the status “Other health worker” (*p*=0.00001) were significantly associated with a very poor knowledge about the novel coronavirus but earning a higher monthly income was significantly associated with a good knowledge of the disease.

In **Table 3**, 27.5%, 95 CI (2.97-2.74) of the participants disagree that their health will be severely damaged if they contract the novel coronavirus. Another 27.5%, 95 CI (3.42-3.16) strongly agree that the novel coronavirus is more infectious than Ebola virus. Only 5.9%, 95 CI (2.34-2.14) strongly agree that they will not go to the hospital, even if they fall ill because of the risk of getting infected with the virus. Some of the participants 28.6%, 95 CI (3.58-3.33) agree that the infection may continue to spread widely in country and in their immediate communities. Only 25.1% of the participants with 95% CI (3.55-3.32) strongly agree that they can protect themselves against being infected. Less than a quarter 22.2%, 95 CI (2.73-2.52) of the participants strongly disagree with the statement that they are more likely to get infected with the virus than other people. However only 25.7%, 95% CI (2.81-2.58) of the participants disagree that receiving a letter or package from abroad can put them at risk of getting infected with the new coronavirus.

**Table 3:** Risk Perceptions of the participants about COVID-19 (n = 510)

| **Risk perception items** | **Responses n (%)** |  |  |  |  |
| --- | --- | --- | --- | --- | --- |
|  | **Strongly disagree** | **Disagree** | **Neutral** | **Agree** | **Strongly agree** |
| My health will be severely damaged if I contract novel Coronavirus. | 99 (19.4) | 140 (27.5) | 81 (15.9) | 117 (22.9) | 73 (14.3) |
| I think novel coronavirus is more contagious than EVD. | 101 (19.8) | 59 (11.6) | 80 (15.7) | 130 (22.5) | 140 (27.5) |
| Even if I fall ill with another disease, I will not go to hospital because of risk of getting Infected | 156 (30.6) | 188 (36.9) | 85 (16.7) | 51 (10) | 30 (5.9) |
| Novel coronavirus may inflict serious damage in my Community. | 82 (16.1) | 49 (9.6) | 90 (17.6) | 132 (25.9) | 157 (30.8) |
| Novel coronavirus may continue To spread widely in the country. | 86 (16.9) | 71 (13.9) | 100 (19.6) | 146 (28.6) | 107 (21) |
| I am more likely to get the novel coronavirus than other people | 113 (22.2) | 133 (26.1) | 139 (27.3) | 83 (16.3) | 42 (8.2) |
| I believe I can protect myself From the novel coronavirus. | 75 (14.7) | 35 (6.9) | 120 (23.5) | 152 (29.8) | 128 (25.1) |
| Receiving a letter or package from Abroad can put me at risk of getting infected with the new Coronavirus | 115 (22.5) | 131 (25.7) | 110 (21.6) | 102 (20) | 52 (10.2) |

**Key:** EVD=Ebola Virus Disease

A *Kruskal Wallis* test in **Table** 4, shows that there was a significant difference of the perceived risk rank across gender (*P*=0.044). Female participants expressed higher risk rank than their male counterpart. The perceived risk rank was also statistically different across the age groups (*P*=0.039) with an increased risk rank as the participant gets older. A post-hoc test showed a statistically significant difference in the perceived risk rank between 55+ and 18-25 age groups (*P*=0.009), 46-55 and 18-25 (*P*=0.018). There was no statistically significant difference in the perceived risk rank between the age groups of 26-35 and18-25 (*P*=0.069), 36-45 and 18-25 (P=0.083), 36-45 and 26-and 36-45 (*P*=0.109) and 55+ as well as 46-55 (*P*=0.442).

**Table 4:** Relationship between risk perceptions and sociodemographic characteristics of the participants about COVID-19 (n = 510)

| **Sociodemographic characteristics** | **Mean ranks** | **P value** |
| --- | --- | --- |
| **Gender** |  |  |
| Male | 246.27 | 0.044* |
| Female | 274.13 |  |
| **Age** |  |  |
| 18 – 25 | 289.59 | 0.039* |
| 26 – 35 | 256.08 |  |
| 36 – 45 | 250.96 |  |
| 46 – 55 | 244.03 |  |
| 55+ | 191.47 |  |
| **Current level of education** |  |  |
| Less than SSCE | 145 | 0.68 |
| SSCE | 253.93 |  |
| Diploma | 222.21 |  |
| Bachelor's degree | 259.19 |  |
| Master's degree | 251.14 |  |
| Professional degree | 272.97 |  |
| Doctorate | 273.03 |  |
| **Region of Residence** |  |  |
| South East | 274.32 | 0.25 |
| North Central | 252.35 |  |
| South-South | 339.53 |  |
| North West | 249.55 |  |
| South West | 239.97 |  |
| North East | 257.59 |  |
| **Monthly income (Naira)** |  |  |
| <30, 000 | 245.80 | 0.48 |
| 30, 000 – 59, 000 | 236.25 |  |
| 60, 000 – 89, 000 | 260.55 |  |
| 90, 000 – 109, 000 | 257.68 |  |
| ≥110, 000 | 268.19 |  |
| **Are you a HCW?** |  |  |
| No | 234.70 | <0.001** |
| Nurse | 311.85 |  |
| Physician | 338.18 |  |
| CHW | 193.41 |  |
| Pharmacist | 306.39 |  |
| MLS | 314.82 |  |
| Other healthcare worker | 237.28 |  |

**KEY:** SSCE= Senior School Certificate Examination; HCW= Healthcare worker; CHW= Community health worker; MLS= Medical Lab Scientist; \*Statistically significant at $p < 0.05$ , 95% CI

Likewise, the *Kruskal Wallis* test showed a significant difference in the perceived risk rank across healthcare workers (*P*=0.001) with Physicians, medical laboratory scientists, pharmacists and nurses showing significantly higher perceived risks than non-health or community health workers.

A Chi square test as shown in **Table 5** revealed that statements like *“Avoided travel to novel coronavirus high risk areas”* (p=0.017), *“Avoided eating outside of the home”* (p=0.001), *“Wore a face mask”* (p=0.01) and *“Ate a balanced diet”* were statistically significant across gender, notwithstanding there were no relationship between gender and statements like, “*washed hands with soap and water*”, “*Avoided touching the eyes, nose, and mouth with unwashed hands*”, “*Covered your cough or sneeze with a tissue, then throw the tissue in the trash*”, “*Avoided close contact with sick people*”, “*Took a supplement*” and “*Disinfected surfaces*”.

**Table 5:**
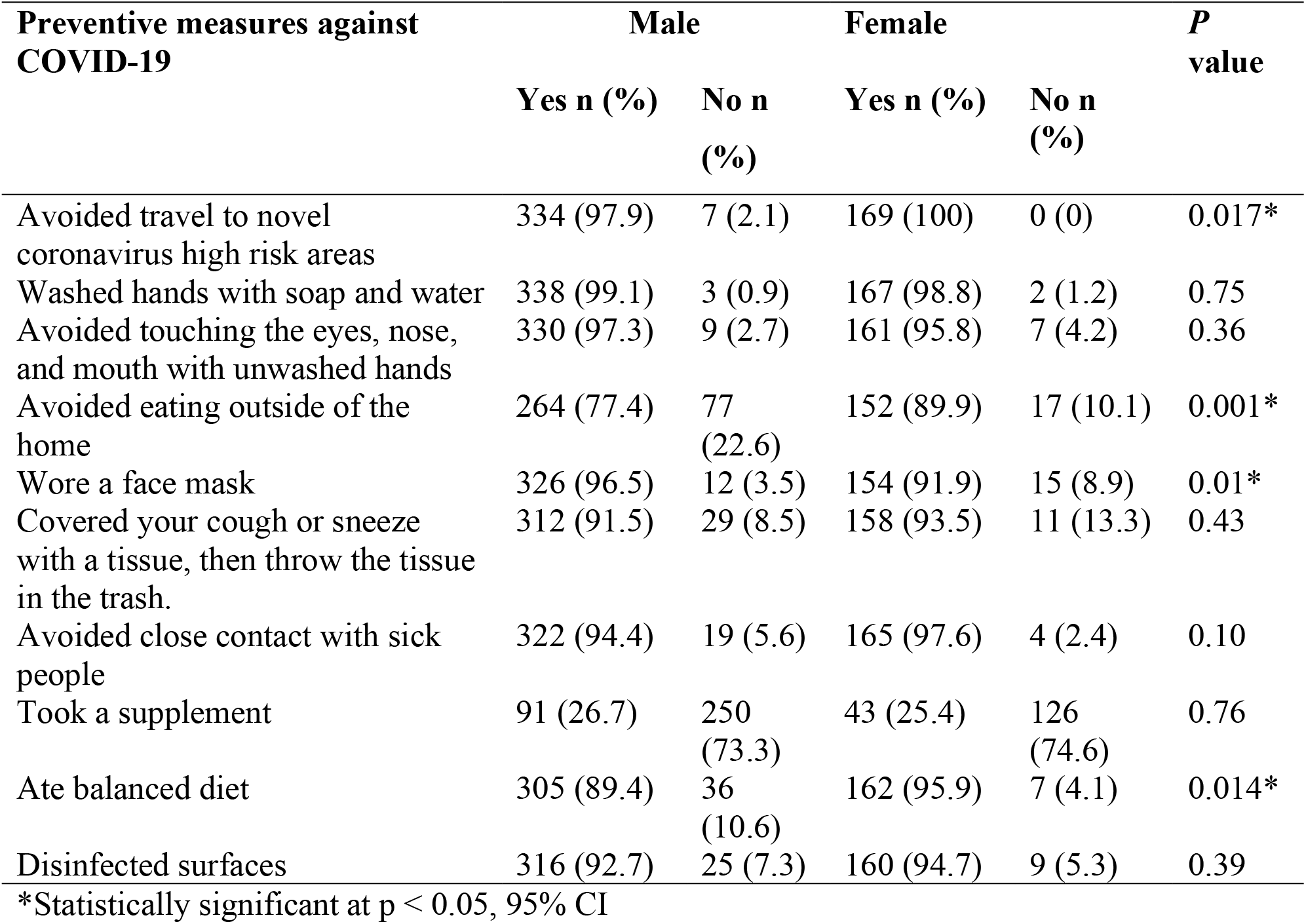
Infection and prevention control measures of participants by gender

## Discussion

We found that a large proportion of the participants (95.9%) were aware of the novel coronavirus pandemic but about half (48%) had a good general knowledge about the disease, its methods of spread, and prevention. This is in agreement with the findings in other studies^14, 15,16^. In like manner, a multinational study in Nigeria and Egypt revealed that good proportion (45.8%) of the participants had a satisfactory knowledge about the disease^17^. Similarly, in another study conducted among the Iranian population, a large proportion of the study population (56.5%) had sufficient knowledge of COVID-19 transmission and symptoms^16^. Traditional media platforms such as newspapers, television and radio, represented the most important sources of information in this study, contrary to another^15^, in which social media platforms, and the internet were more patronized. Indeed, research shows that public engagement with spurious information is greater than with legitimate news from mainstream sources, making social media a powerful channel for propaganda^18^. Fake news on social media about potential drugs, including chloroquine has led to the shortage of this medicine because of the high demand making patients who actually need them to be out of the medicines^19^.

From our study, level of education influences the knowledge of COVID-19 in such a way those with higher degrees tend to have a better knowledge of the disease when compared to those with lower qualifications. Likewise, those with higher monthly income have a better knowledge of the disease when compared with lower monthly income. This is consistent with other studies ^20^. This may be as a result of lack of access to credible and timely information about the virus for poorly educated citizens. Being a healthcare worker was significantly associated with a poor knowledge of the disease. This is not consistent with cross-sectional, web-based studies conducted among health care workers, where it was reported that healthcare workers had good knowledge of COVID-19^21, 22, 23^. Other studies reported that being a healthcare worker or having a background medical knowledge was associated with a good knowledge about the disease^17,24^. The knowledge of healthcare workers cannot be over emphasized in a pandemic like COVID-19, knowing fully well that scientists are still studying the novel coronavirus.

Participants reported high risk perception, this is in concert with findings in a study^23^. Notwithstanding, a contrary study found a relatively low risk perception among U.S citizens^8^. Most participants reported that their health will not be negatively affected even if they contract the virus but they were concerned that the virus may continue to spread in the country. However, a low proportion (25.1%) of our participants believed that they can protect themselves against the virus. The fear of getting infected with the virus when seeking medical care in the hospital was very low (5.9%). Likewise, the participants did not perceive that receiving letters or package abroad can pose a risk of infecting them, which is in agreement with findings from a study in the U.S and U.K that receiving a package from overseas did not pose a greater risk of infection with the virus^9^.

There was a significant difference across gender with the females having a higher risk perception than the males. This is in agreement with another study^25^. However, the male gender was found to have a high-risk perception towards the virus according to another study^26^. The younger adults have the highest risk perception towards the virus as well. The perceived risk of working as a healthcare worker also differ significantly. This is similar to the findings of a study^22^, in which healthcare workers have a higher risk perception rank than the general population because of their close contact with suspected/confirmed COVID-19 cases.

Avoiding travel to high risk areas was significantly different across gender in which females were much less likely to travel to areas with high cases. Likewise, avoiding eating out significantly contrasted with gender in which the female gender was less likely to eat out than their male counterpart. Wearing of face mask and eating balanced diet also significantly differ across gender in which males were more likely to wear face masks but females are more likely to eat balanced diet. Other Studies^27, 28^, revealed that diet and nutrition invariably influence the immune system competence to fight infections and determine the risk and severity of infections. Improving the diet quality in susceptible individuals for COVID-19 might alleviate their risk of severe infection. Nigeria Centre for Disease Control (NCDC), the Nigerian public health institute offers infection prevention and control measures to healthcare workers as well as the general public^29^. The awareness and sensitization campaigns by the Federal Ministry of Health (FMoH) and the NCDC have reflected in the way participants practice preventive measures against the novel coronavirus.

Lastly, more than half of the participants selected a health care–seeking option that could lead to reduction in transmission of SARS-CoV-2. This is consistent with another study^9^, in the U.S and the U.K in which just one-fourth of the participants chose health care seeking responses that could lead to increase in the transmission of the novel coronavirus. Thus, clear messaging on the recommended care-seeking action by the NCDC has really helped in informing the general public about the common symptoms of COVID-19 and how to seek medical care.

## Limitations

The distribution of the survey through the internet allowed only those who can read and have internet access to participate and likewise the distribution of responses by participant’s regions may differ from the general population owing to the fact that samples from South-South and the South-East were small. Another limitation could be that our data was skewed to the young adults. Also lack of inclusion of those with chronic illness in this study was also a limitation as novel coronavirus tends to be more deleterious on those with chronic diseases.

## Conclusion

In general, our participants had a good knowledge of COVID-19 with a low risk perception among non-healthcare workers but a high-risk perception of getting infected with the novel coronavirus was observed among healthcare workers. This knowledge is mainly acquired through the traditional media platforms. However, knowledge was lower among less educated and lower income groups. Interventions may require more efforts or using different methods to communicate with these groups.

## Data Availability

Data will be made available on request.

## Acknowledgement

We would like to thank Dr. Musa M. Watila (NIHR University College London Hospitals Biomedical Research Centre, UCL Institute of Neurology, London) and Dr. Roland N. Okoro (University of Maiduguri) for their insights. Our participants are highly appreciated for taking part in this survey.

